# Psychological flexibility and the moderating role of the therapeutic working alliance in Acceptance and Commitment Therapy in Daily Life (ACT-DL) in an early psychosis sample

**DOI:** 10.1101/2022.01.19.22269524

**Authors:** Evelyne van Aubel, Thomas Vaessen, Ruud van Winkel, Ginette Lafit, Annelie Beijer-Klippel, Wolfgang Viechtbauer, Tim Batink, Mark van der Gaag, Therese van Amelsvoort, Machteld Marcelis, Frederike Schirmbeck, Lieuwe de Haan, Ulrich Reininghaus, Inez Myin-Germeys

## Abstract

**Background:** We investigated treatment effects of Acceptance and Commitment Therapy in Daily Life (ACT-DL) on psychological flexibility (PF) and the moderating role of the therapeutic working alliance on these effects in patients with early psychosis.

**Methods:** ACT-DL is an ecological momentary intervention (EMI) combining face-to-face ACT with a smartphone app. In the multi-center INTERACT randomized controlled trial, n=148 early psychosis individuals were randomized to either treatment as usual (TAU as the control condition, n=77) or to ACT-DL in addition to TAU (ACT-DL + TAU as the experimental condition, n=71). We assessed global PF and the therapeutic alliance with self-report questionnaires. In addition, we used the experience sampling methodology (ESM) to assess PF with a momentary (in-the-moment and since-the-previous-beep openness) and an evening (daily PF) questionnaire. Assessments took place at baseline, post-intervention (POST), six (FU6), and twelve months (FU12) follow-up.

**Results:** Global (B=19.49 to 33.14; all P-values<.001) and daily PF (B=0.68; P-value<.001) improved equally in both conditions at each time point. Individuals in the ACT-DL condition improved more than those in TAU on momentary openness (in-the-moment openness at POST (B=0.32; P-value=0.007) and since-the-previous-beep openness at POST (B=0.33; P<.001) and FU6 (B=0.23; P-value=0.025). Client-perceived working alliance moderated in-the-moment openness such that larger improvements in openness at POST (B=0.05; P-value<.001) were found in ACT-DL in individuals with higher working alliance scores.

**Conclusion:** Our results provide partial support for the capability of ACT-DL to improve daily life measures of openness, and emphasize the importance of the therapeutic relationship in supporting processes of change.

## 1 Introduction

Psychological inflexibility, defined as the rigid dominance of psychological reactions over chosen values and contingencies in guiding action (Bond et al., 2011), has been associated with poor mental health in various populations and clinical diagnoses (Chawla & Ostafin, 2007; Hayes et al., 2006; Kashdan & Rottenberg, 2010; Levin et al., 2013). Psychological inflexibility has also been linked to psychotic experiences and associated distress (Pankey & Hayes, 2003), and may therefore be an interesting psychotherapeutic target in the treatment of psychosis (Pankey & Hayes, 2003).

Psychological inflexibility consists of various sub-processes of which two have received the most attention in the psychosis literature. First, experiential avoidance, defined as those strategies to avoid painful thoughts, emotions, or sensations, has been associated with higher levels of depression, anxiety, and distress in individuals with a psychotic disorder (Morris et al., 2014; Perry et al., 2011; White et al., 2013). Second, cognitive fusion, defined as the tendency to see thoughts as reality, has been found to play a role in aggravating delusions, auditory hallucinations, and their associated impact and distress (Goldstone et al., 2011; Morris et al., 2014; Peters et al., 2012; Varese et al., 2016). Studies making use of the Experience Sampling Method (ESM), a technique to assess symptoms, emotions, and their context multiple times a day in the flow of daily life (Csikszentmihalyi & Larson, 1987; Myin-Germeys et al., 2018) found that psychological inflexibility and experiential avoidance more specifically are not only maladaptive at the trait level, but also at the state level, when used in daily life as a coping strategy to deal with difficult experiences. In this respect, two ESM studies showed that momentary experiential avoidance is associated with momentary paranoia, both concurrently and at the next time point, in college students (Udachina et al., 2009) as well as in individuals with a psychotic disorder (Udachina et al., 2014).

*Acceptance and Commitment Therapy* (ACT) is a third wave behavioral therapy that aims to increase *psychological flexibility* (PF) (Hayes et al., 2006). For individuals with a psychotic disorder, this means that they will learn to take a more open, aware, and active stance towards aversive experiences (Pankey & Hayes, 2003). In turn, an improvement in these processes is expected to positively affect various outcomes including symptom-related distress and functioning. This may also be relevant for individuals in the early stages of psychosis, including individuals at *ultra-high-risk* (UHR) for psychosis (Yung et al., 2005)^a^ and individuals with a *first episode of psychosis* (FEP) (Fusar-Poli et al., 2013; Linscott & Van Os, 2013; Van Os & Linscott, 2012; Van Os & Reininghaus, 2016), given that these individuals suffer from high levels of psychotic symptom distress (Birchwood, 2003; Rapado-Castro et al., 2015; Rekhi et al., 2019; Steel et al., 2007; R. S. Wilson et al., 2020), and struggle with negative symptoms and related impairments in functioning and quality of life (Devoe et al., 2020; Fusar-Poli et al., 2015; Schlosser et al., 2014).

However, only few clinical trials of ACT for psychosis have looked into treatment effects on PF and its related sub-processes, and have only looked into trait outcomes measured with global self-report questionnaires. While some studies found significant effects favoring ACT over TAU on acceptance (Gumley et al., 2017; Spidel et al., 2018) and mindfulness (Gumley et al., 2017; White et al., 2011), other trials did not find larger differences between conditions on acceptance (Shawyer et al., 2012, 2017; White et al., 2011). In more recent studies, ACT has been offered in a blended care approach (Batink et al., 2016; Levin et al., 2017; Ly et al., 2014) combining face-to-face ACT sessions with an *Ecological Momentary Intervention* (EMI). EMIs deliver real-time momentary psychological interventions in daily life, tailored to what individuals need in a given context, making use of mobile technologies including smartphone *applications* (apps) or other mobile devices (Heron & Smyth, 2010; Myin-Germeys et al., 2016). To our knowledge, only one study investigated treatment effects of an ACT-based EMI (ACT Daily) on daily life PF in a small sample of individuals with depression and/or anxiety who were following ACT treatment. Interestingly, this study found significant improvements after 2 weeks use of the EMI on momentary acceptance, defusion, valued living, and mindfulness as measured in daily life (Levin et al., 2017).

In our group, we have developed the ACT in Daily Life intervention, a blended care intervention combining face-to-face ACT sessions with an EMI delivered via a mobile application (the ACT-DL app) (Vaessen et al., 2019). It is expected that ACT-DL will optimize treatment effects by capitalizing on the benefits of both EMIs and of standard face-to-face psychotherapy. As for the ACT-DL app, individuals receive notifications with short questionnaires containing items on their current mood, symptoms, and context to increase mindful awareness, followed by a visual cue of an ACT metaphor or a short ACT exercise. The use of the app may therefore facilitate the real-world acquisition and generalization of new coping skills learned within therapy, which may eventually lead to sustainable changes in intended outcomes under real-world (ecological) conditions (Reininghaus et al., 2016).

As to the face-to-face sessions, they allow patients to benefit from the working alliance they can build with their psychotherapist. Indeed, the therapeutic working alliance has been identified as a predictor of treatment engagement and outcome (Bourke et al., 2021; Browne et al., 2021; Shattock et al., 2018), as well as a moderator of improvement in clinical and functional outcomes (Chen et al., 2018; Kay-Lambkin et al., 2017). However, no research to date has investigated the moderating role of the alliance on ACT processes measures. Investigating the role of the therapeutic alliance may be especially important within ACT, given that the therapist needs to be aware of and function as a role model of these PF processes within the session itself (Vilardaga & Hayes, 2009; K. G. Wilson & Sandoz, 2010). These therapeutic qualities are needed to avoid therapeutic ruptures or impediments of therapeutic processes to unfold (for a case example, please see (Walser & O’Connell, 2021)). For that reason, we believe that a good therapeutic alliance may support the improvement of PF.

As such, in the current paper, we will evaluate whether ACT-DL improves psychological flexibility both measured with a global self-report questionnaire as well as in daily life. In addition, we will evaluate whether the therapeutic working alliance moderates this improvement. We hypothesize that:

(1) ACT-DL, in comparison to TAU, will lead to larger global and daily life improvements in PF at post-intervention, 6-month, and 12-month follow-up.
(2) Improvements in global and daily life PF will be moderated by the client-perceived and/or the therapist-perceived working alliance, such that those participating in ACT-DL with better alliances will improve most on these processes.

## 2 Methods

### 2.1 Study design and participants

In the multi-center INTERACT *randomized controlled trial* (RCT) (Dutch Trial Register, ID: NTR4252), participants were recruited from secondary mental health services at clinical sites within five regions in The Netherlands and Belgium. Inclusion criteria were: 1) aged 15-65, 2) UHR (without prior use of antipsychotic medication) or FEP (onset within last 3 years) as assessed by the Comprehensive Assessment of an At Risk Mental State (CAARMS) (Yung et al., 2005) or Nottingham Onset Schedule (NOS) (Singh et al., 2005), and 3) sufficient command of the Dutch language. Exclusion criteria were: 1) a primary diagnosis of alcohol/substance abuse and dependence, assessed with the Mini-International Neuropsychiatric Interview (MINI) (Sheehan et al., 1998), and 2) severe endocrine, cardiovascular, or brain disease. After informed consent was signed and baseline assessment finalized, participants were randomly allocated (50:50) to a control condition of *treatment as usual* (TAU) or to an experimental condition of ACT-DL in addition to TAU (ACT-DL+TAU). The trial received ethical approval from the *Medical Ethics Review Committees* (MERC) at Maastricht University Medical Centre (MUMC), the Netherlands (reference: NL46439.068.13) and the University Clinic Leuven, Belgium (reference: B322201629214). The full study methodology is detailed in the study protocol (Reininghaus et al., 2019).

### 2.2 Interventions

Participants allocated to TAU continued to receive all the treatment they received prior to the start of the study, which we assessed using a study-specific service-use checklist. TAU consisted of good standard care delivered according to local and national service guidelines and protocols, and included cognitive behavioral therapy for psychosis (CBTp) in some sites.

Participants allocated to ACT-DL received, in addition to TAU, eight weekly individual face-to-face ACT sessions with a trained ACT clinician, including one psycho-education session followed by a session on a new ACT component every week. In a final session, all previous components were integrated into the topic of PF. Participants used the ACT-DL EMI in between the therapy sessions. The EMI was delivered via the PsyMate™ (www.psymate.eu) app (hereafter: the ACT-DL app), which participants were instructed to use for three subsequent days starting from the first day after therapy had taken place. During these three days, participants received eight notifications (‘beeps’) a day, prompting them to fill in a short ESM questionnaire with various items on mood, symptoms, and their current context to help increase present awareness. Every beep questionnaire was followed by either an ACT-exercise or a visual cue of an ACT-metaphor (50:50 ratio). Finally, participants could initiate an on-demand ACT exercise whenever they were struggling with difficult thoughts or emotions. Please see Vaessen et al. (2019) for a more detailed description of the intervention.

### 2.3 Measures and procedures

#### 2.3.1 Retrospective measures

**Global psychological flexibility** and its related sub-processes were measured with the self-reported Flexibility Index Test 60 (FIT-60) (Batink et al., 2012; Batink & Delespaul, 2015) total score and the scores on its subscales (i.e., acceptance, defusion, mindfulness, self as context, values, and committed action) (Cronbach’s α between .70 and .89) at baseline, post-intervention, 6-, and 12-month follow-up. The FIT-60 measures PF on a trait level, asking participants to reflect on how they behave in general.

**Therapeutic Working Alliance** was assessed with the self-reported Working Alliance Inventory – Short Form (WAI-SF) (Hatcher & Gillaspy, 2006; Horvath & Greenberg, 1989) at post-intervention. Whereas the client-rated version asked clients to reflect on the contacts they had with their ACT therapist (in ACT-DL) or with the clinician who they saw and talked to most frequently (in TAU), the therapist-rated version asked either the ACT therapist or the main clinician to reflect on their working alliance with the participant. In the current study, we used the total WAI-SF scale (Cronbach’s α=.92 for client-perceived and α=.87 for therapist-perceived alliance).

**Symptom severity** was assessed at baseline with the total score of the Brief Psychiatric Rating Scale (BPRS) (Ventura et al., 1993), reflecting the previous two weeks before the interview (Cronbach’s α=.67). We controlled for baseline symptom severity in all analyses given persistent findings of associations with lower levels of PF (Chawla & Ostafin, 2007; Kashdan et al., 2006; Kashdan & Rottenberg, 2010; Levin et al., 2013).

#### 2.3.2 Daily life ESM measures

The Experience Sampling Method (ESM) (Csikszentmihalyi & Larson, 1987; Myin-Germeys et al., 2018) is a structured, random time-sampling diary technique to measure moment-to-moment as well as daily variability in several ACT psychological flexibility processes, in the real world and in real time, with high ecological validity. ESM measures were assessed at baseline, post-intervention, and 6-month follow-up.

During each ESM assessment period, participants received 10 notifications per day during a period of six consecutive days on the PsyMate™ app, making use of a stratified random sampling technique with ESM assessments scheduled at random within set blocks of time between 7:30 AM and 10:30 PM. In addition, participants were asked to self-initiate an evening questionnaire that was available between 8:00 PM and 11:00 PM.

### Momentary openness

Each ESM questionnaire prompted participants with Likert scale items (1: not at all; 7: very much) on how open (versus experientially avoidant) they were towards difficult experiences. We operationalized **in-the-moment openness** with the item “*I wish I could get rid of my negative feelings (reverse scored)*”. Second, **since-the-previous-beep openness** was operationalized averaging the two items “*I tried not to think about [negative emotions or thoughts] (reverse scored)*” and “*I let my thoughts be without reacting to them*” when individuals responded >2 on the item “*Since the last beep, I’ve had negative emotions or thoughts*”, and set to missing if <3, discarding these observations.

### Daily PF

Daily PF was operationalized as the average of five items on the evening questionnaire (scored on 7-point Likert scale - 1: not at all; 7: very much) including items on open (“*Today, I could let go of my negative feelings without acting upon them*”, “*In general I have ruminated a lot today (reverse scored)*”), aware (“*Today, I was consciously aware of the activities that I did*”), and engaged (“*Today, I have invested in what I find important in my life*”) processes. Please refer to Figure 1 for more details on the timing of the various daily life ACT processes.

**Figure 1.**
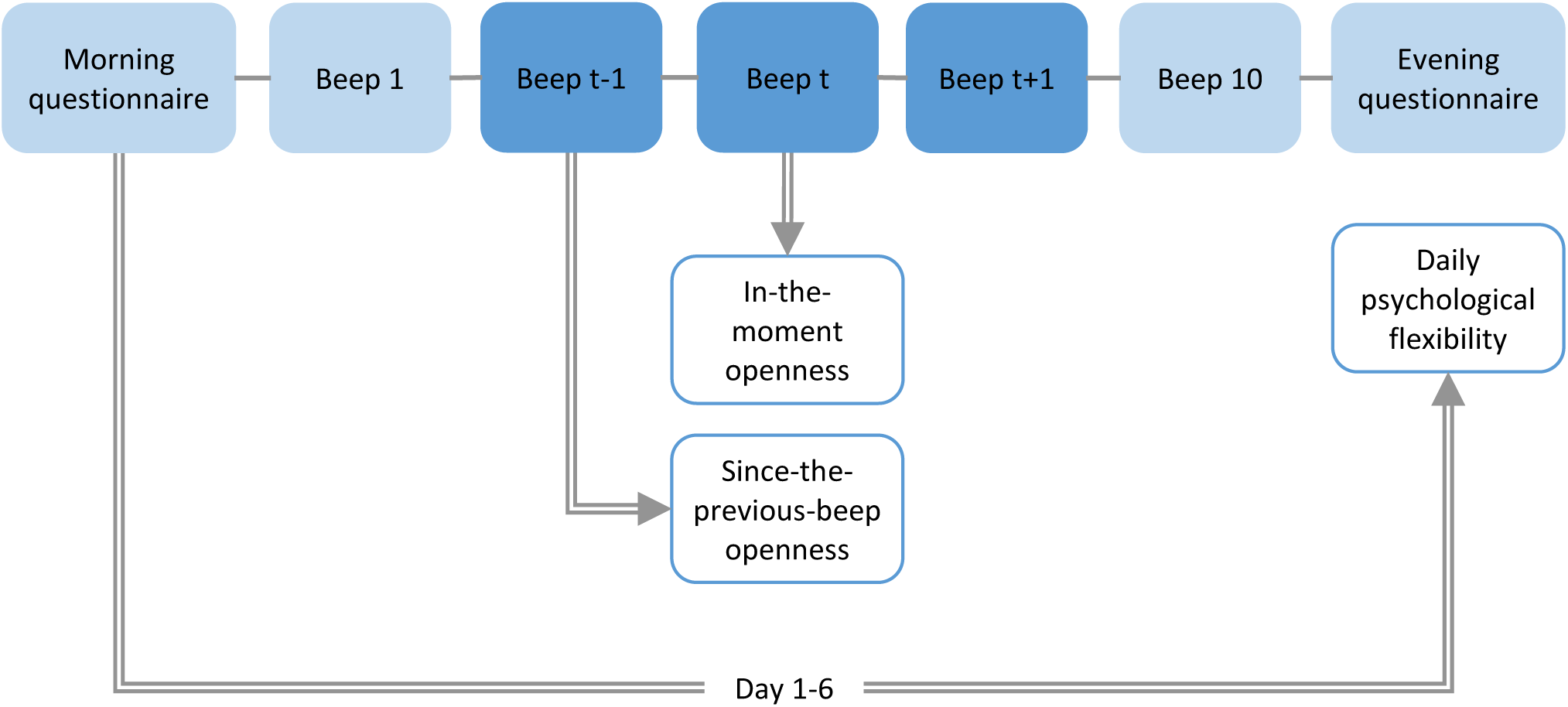
Timing of the various daily life psychological flexibility variables. The double arrow represents the ESM assessment window. Momentary and since-the-previous-beep openness are both assessed at beep t. In-the-moment openness reflects the moment right before the beep went off. Since-the-previous-beep openness reflects the time since the previous beep. Daily PF assessed the experience of the previous day.

We reported information on ESM compliance and its psychometric properties in Supplement 1 and Supplementary Table 1.

### 2.4 Statistical analysis

#### 2.4.1 Changes in psychological flexibility

Differences in treatment effects over time on global, momentary, and daily psychological flexibility were tested with separate repeated measures mixed-effects models using the MIXED command in Stata (StataCorp, 2015), with the FIT-60 total score, in-the-moment openness, since-the-previous-beep openness, and daily psychological flexibility as the dependent variable. Independent variables in these models were time (as a four-level factor, for baseline, post-intervention, and the 6- and 12-month follow-ups), condition (dichotomous: ACT-DL + TAU vs. TAU only), and the time × condition interaction. We controlled for region (as a five-level factor) and group status (dichotomous: UHR vs. FEP) in all models, as well as for baseline BPRS scores by including them as covariates in the models. In the analysis of the FIT-60 total scores, we took into account within-subject clustering of the repeated measures by allowing the models’ level-1 residuals to be correlated making use of a completely unstructured error variance-covariance matrix. In our models with ESM outcomes, multilevel mixed-effects models were used with multiple ESM observations (level-1) nested within time points (now as a three-level factor, for baseline, post-intervention, and the 6-month follow-up) (level-2) and time points nested within subjects (level-3). We added level-3 random intercepts to the models. In addition, for the errors at level 1, we assumed an autoregressive error variance-covariance matrix of the exponential type for momentary measures and of the autoregressive type for daily measures, which allowed our models to account for unequally spaced time values in the ESM data.

In addition to our model on global PF, we used the nlme package (Pinheiro et al., 2021) in R (RStudio Team, 2015) to fit a related outcomes model, with all FIT-60 subscales as six dependent variables in one model. The independent variables were the same as the ones described above. This model included a random intercept for each outcome and assumed that the residual errors of each outcome were multivariate normally distributed and uncorrelated over time. Given the assumption that the six ACT sub-processes were not independent from each other, all variances of and co-variances between random intercepts (one for each dependent variable within subjects), as well as of and between the residual errors of these outcomes were estimated. This model was similar to our univariate model described above, except that all six outcomes were measured simultaneously rather than sequentially, which makes comparison of the sub-scales possible (Baldwin et al., 2014).

Within our univariate models, we used the TEST command in Stata to perform a joint omnibus test of no difference between the two conditions at all three time points after baseline (Wald-type test with df=3 and α=.05), and if statistically significant, at each time point separately (each tested at α=.05). The same command was used to assess the main effect of time at all three time points simultaneously. To aid interpretability of our findings, we calculated the predicted margins, SE, and estimated differences between conditions for these outcomes using the MARGINS and the CONTRAST command respectively.

Within our multivariate model, the linearHypothesis command using the car package (Fox & Weisberg, 2019) in R was used to perform these same tests for each ACT process individually. To date, it is not possible to calculate predicted margins in R after having fit a related outcomes model. We therefore based the margins for the FIT-60 subscales on six univariate repeated measures mixed-effects models on these outcomes, as described for the FIT-60 total score (above). These predicted margins therefore need to be interpreted with caution because they do not reflect the post estimations of the joint multivariate model.

#### 2.4.2 Therapeutic working alliance as a moderator

To test whether the therapeutic working alliance moderated treatment effects on psychological flexibility, we added client-perceived and therapist-perceived WAI-SF total scores (separately) as moderators to the repeated measures mixed-effects models outlined above. Within each model, this meant that we added the condition × working alliance, the time × working alliance interaction, and finally the condition × time × working alliance interaction.

After each model fit, we performed a joint omnibus test over all time points of the three-way interaction effects with the TEST command, followed by time-specific contrasts if statistically significant at α=0.05. All models were fitted using *restricted maximum likelihood estimation* (REML). For the full statistical analysis plan, please refer to our post-registration report^b^.

## 3 Results

The final sample included 70 participants with UHR and 78 with FEP, and was nearly equally divided between men and women (49% for the latter). Participants had a mean age of 25.1 (SD=5.9, range 15 to 47). Detailed information on the INTERACT sample can be found in the study’s main outcome paper (Myin-Germeys et al., 2021). Descriptive statistics for baseline FIT-60, WAI-SF, BPRS symptom severity scores, and ESM measures are reported in Table 1. For more details on data availability in the current study, please refer to Supplementary Figure 1.

**Table 1.** Baseline descriptive mean (SD) scores.

| Table 1. Baseline descriptive mean (SD) scores. |  |  |  |  |  |  |  |  |  |  |  |  |
| --- | --- | --- | --- | --- | --- | --- | --- | --- | --- | --- | --- | --- |
| Measure | N | Total sample (n=148) |  |  | N | ACT-DL+TAU (n=71) |  |  | N | TAU only (n=77) |  |  |
|  |  | Mean | SD (_b; _w) |  |  | Mean | SD (_b; _w) |  |  | Mean | SD (_b; _w) |  |
| Global PF |  |  |  |  |  |  |  |  |  |  |  |  |
| FIT-60 (total) |  | 170.0 |  |  |  |  |  |  |  |  |  |  |
|  | 140 | 7 | 50.67 |  | 66 | 172.24 | 52.56 |  | 74 | 168.14 | 49.20 |  |
| FIT-60 (acceptance) | 139 | 25.48 | 11.71 |  | 66 | 24.72 | 11.69 |  | 73 | 26.18 | 11.76 |  |
| FIT-60 (defusion) | 139 | 22.43 | 13.41 |  | 66 | 21.90 | 13.51 |  | 73 | 22.90 | 13.39 |  |
| FIT-60 (self as context) | 140 | 25.07 | 9.11 |  | 66 | 24.76 | 9.29 |  | 74 | 25.35 | 9.00 |  |
| FIT-60 (here and now) | 139 | 28.05 | 10.49 |  | 66 | 28.66 | 10.78 |  | 73 | 27.51 | 10.25 |  |
| FIT-60 (values) | 140 | 37.33 | 10.19 |  | 66 | 38.61 | 10.23 |  | 74 | 36.19 | 10.09 |  |
| FIT-60 (committed action) | 139 | 31.67 | 11.71 |  | 65 | 33.89 | 11.25 |  | 74 | 29.71 | 11.83 |  |
| Momentary and daily PF |  |  |  |  |  |  |  |  |  |  |  |  |
| In-the-moment openness | 147 | 4.10 | 1.83 | 1.34 | 71 | 4.17 | 1.83 | 1.35 | 76 | 4.03 | 1.84 | 1.33 |
| Since-the-previous-beep |  |  |  |  |  |  |  |  |  |  |  |  |
| openness | 135 | 3.83 | 0.62 | 0.82 | 64 | 3.92 | 0.67 | 0.87 | 71 | 3.75 | 0.57 | 0.77 |
| Daily PF | 126 | 4.22 | 1.04 | 0.82 | 63 | 4.14 | 0.88 | 0.82 | 63 | 4.29 | 1.17 | 0.82 |
| Working alliance |  |  |  |  |  |  |  |  |  |  |  |  |
| WAI-SF (client) | 94 | 43.14 | 9.90 |  | 46 | 43.25 | 10.42 |  | 48 | 43.03 | 9.49 |  |
| WAI-SF (therapist) | 68 | 38.15 | 5.53 |  | 41 | 38.02 | 5.21 |  | 27 | 38.33 | 6.10 |  |
| Symptom severity |  |  |  |  |  |  |  |  |  |  |  |  |
| BPRS (total) | 148 | 40.81 | 8.43 |  | 71 | 39.50 | 7.76 |  | 77 | 42.02 | 8.88 |  |
| Notes. Data represent baseline descriptive mean (SD) scores of trait psychological flexibility (FIT-60 scores), perceived working alliance (WAI-SF), daily life (ESM) measures, and symptom severity (BPRS total scores). For ESM outcomes, we report both the standard deviation between (SD_b) and the standard deviation within (SD_w). |  |  |  |  |  |  |  |  |  |  |  |  |

### 3.1 Changes in psychological flexibility

#### 3.1.1 Global psychological flexibility

As can be seen in Table 2 and Figure 2, the repeated measures mixed-effects model on the FIT-60 total score did not support our hypothesis of increased scores in the ACT-DL+TAU compared to the TAU condition. Yet, the main effect of time was significant (P-value <.001) and time-specific contrasts showed increased mean levels of FIT-60 total scores in comparison to baseline for both conditions at post-intervention (P-value <.001), 6-month (P-value <.001), and 12-month follow-up (P-value <.001).

**Figure 2.**
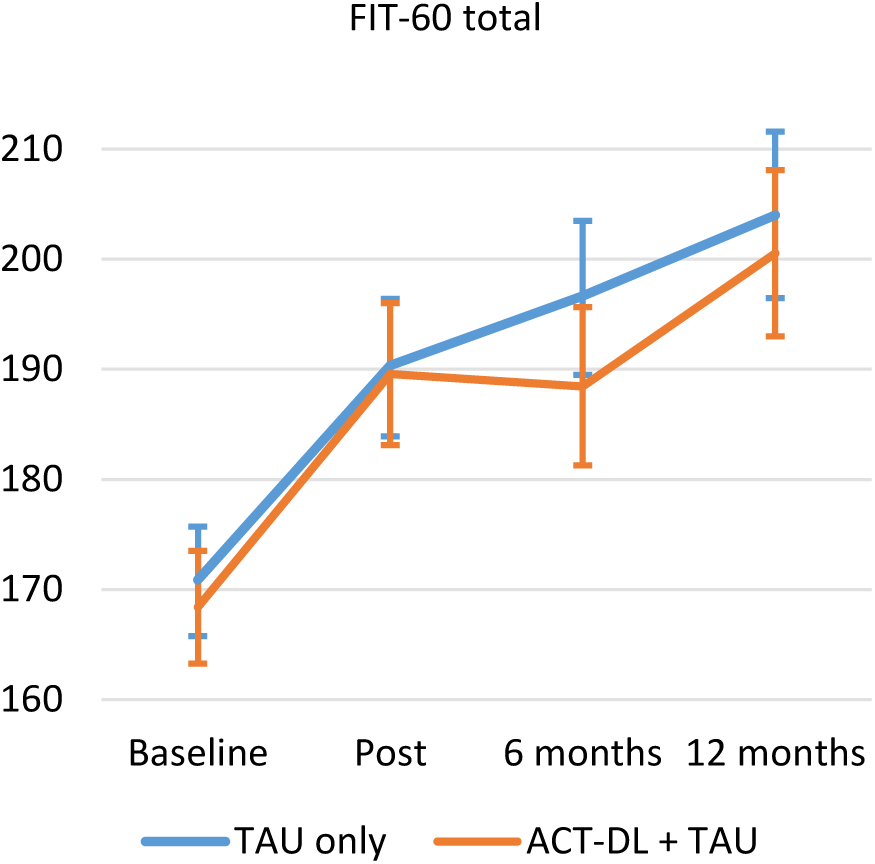
Global psychological flexibility at baseline, post-intervention, 6-, and 12-month follow-up.

**Table 2.**
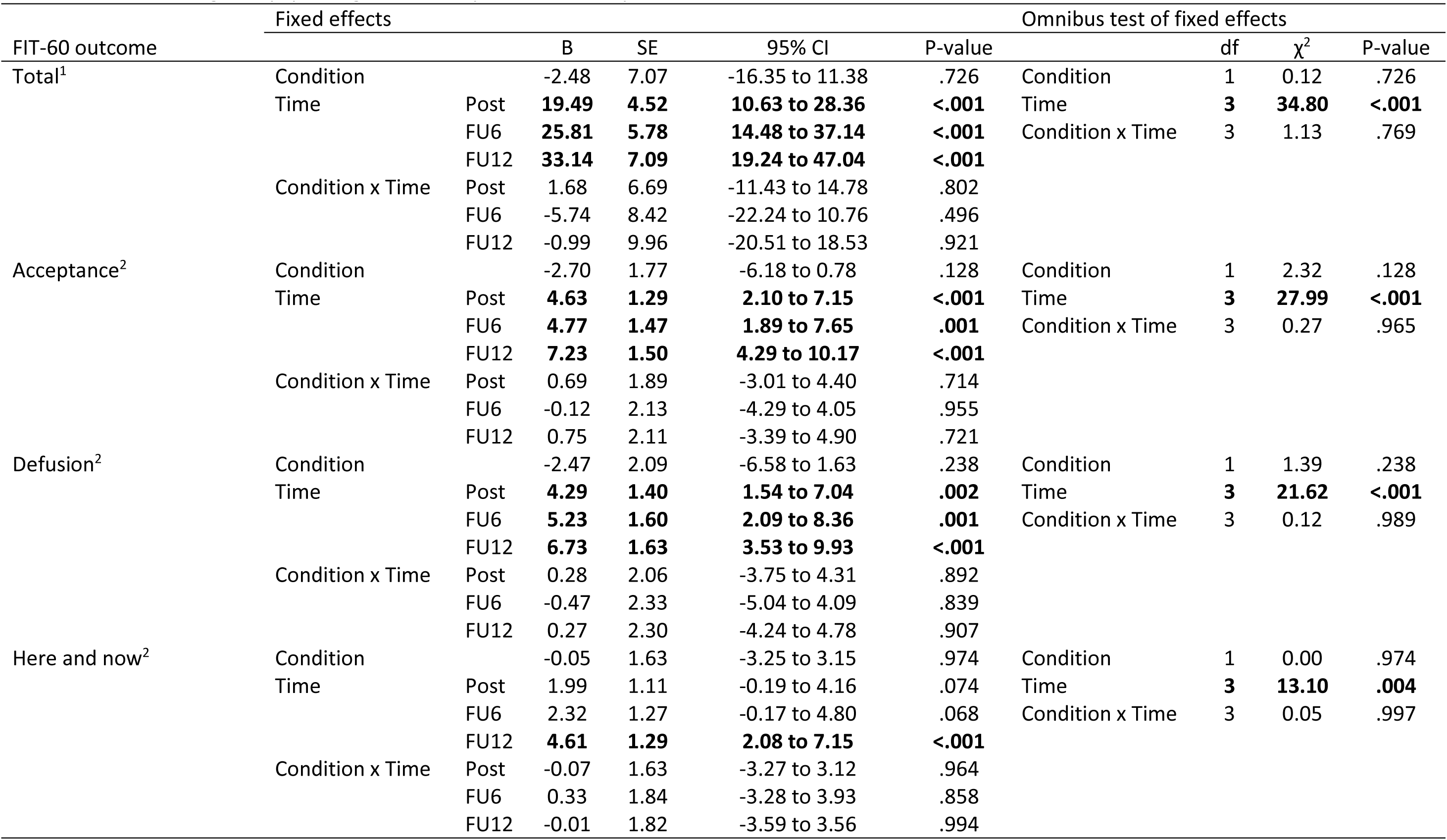
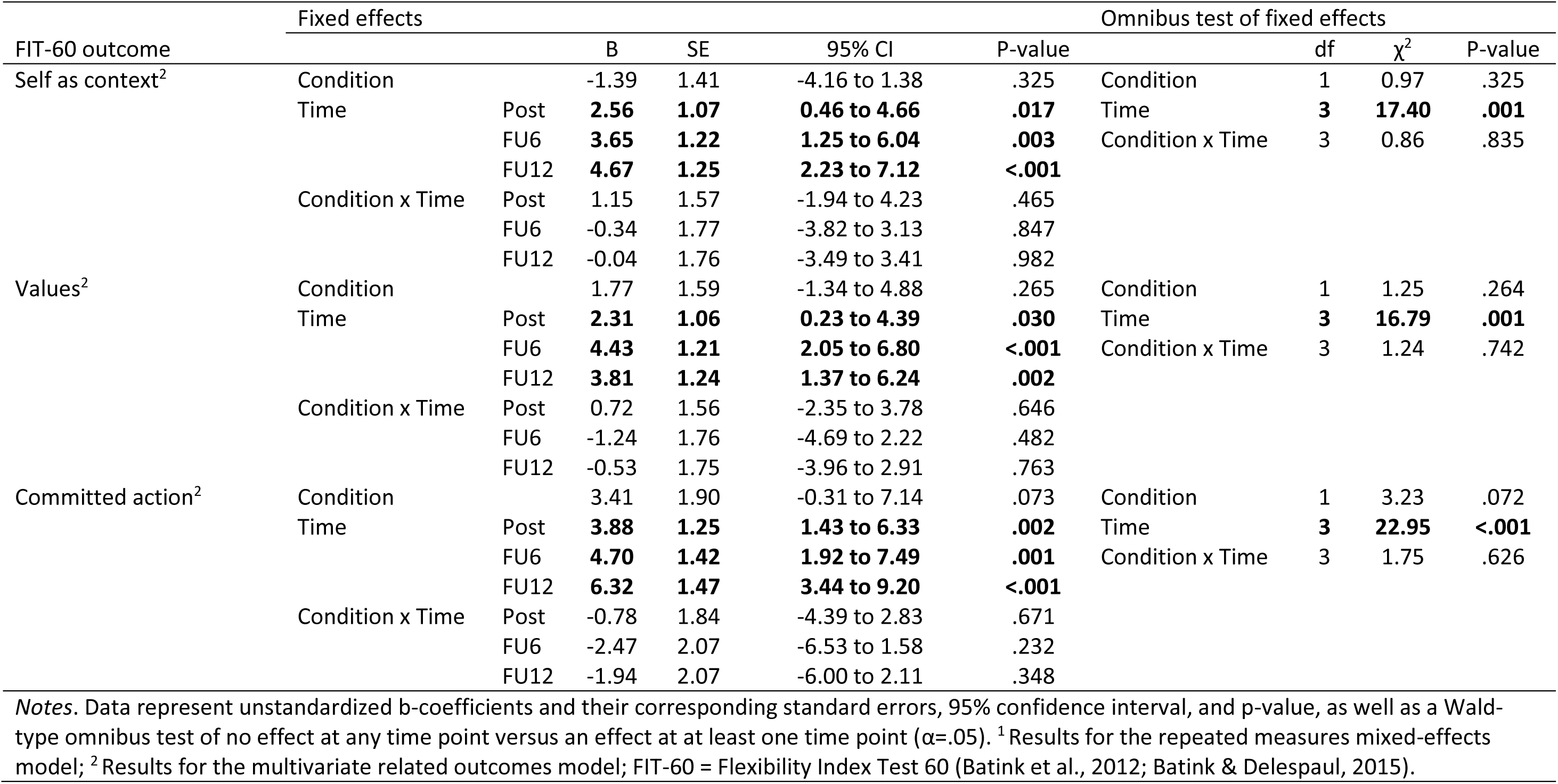
Results of a repeated measures mixed-effects model (FIT-60 total score) and a multivariate related outcomes model (FIT-60 subscale scores) of treatment effects on global psychological flexibility and its six sub-processes.

The results of our multivariate related outcomes model were similar. While there was no support for our hypothesis of increased FIT-60 scores in ACT-DL+TAU compared to TAU on any subscale, we found main effects of time for every subscale (P-values all <.01), including acceptance, defusion, here and now, self as context, values, and committed action (see Table 2). These results suggest that, in both conditions, in comparison to baseline, all ACT skills significantly increased over time in both conditions (P-values all <.05), and did so at each follow-up time point, except for changes in here and now scores, which only reached significance at the 12-month follow-up (P-value<.001).

Similarly, when we plotted the predicted scores on ACT hexaflex radar charts and compared them to norm scores based on a clinical population with heterogeneous psychopathology (Batink & Delespaul, 2015) (Figure 3), we found that FIT-60 scores transitioned from the lower-average range at baseline to the average-upper range at the 12-month follow-up, which may suggest clinically relevant changes in both conditions.

**Figure 3.**
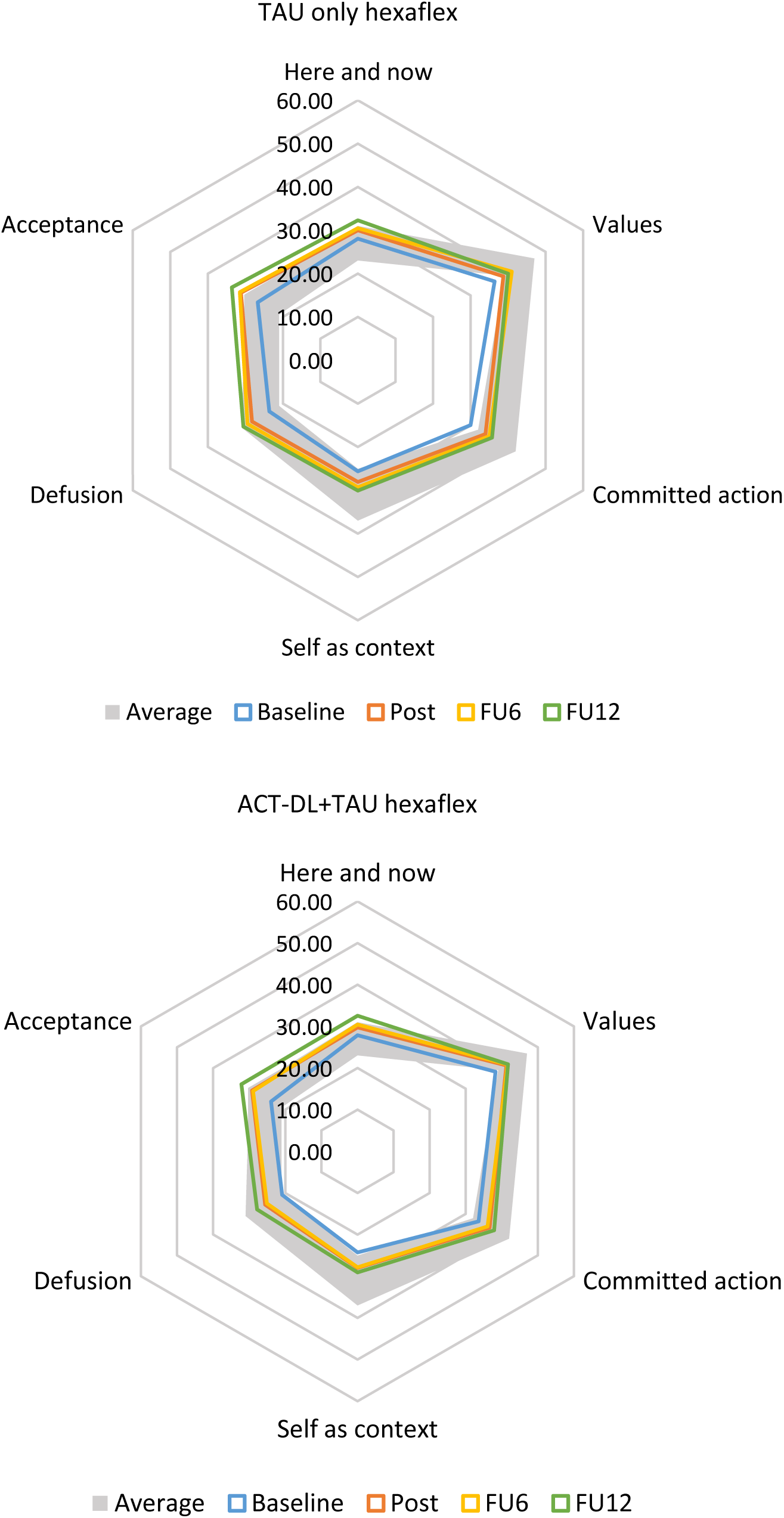
ACT hexaflex scores in TAU only (top) and ACT-DL+TAU (bottom) at baseline, post-intervention, and 6-, and 12-month follow-up. The gray areas represent average norm scores for a clinical population sample (Batink & Delespaul, 2015).

Finally, in line with the lack of significant Condition x Time interactions in both models, simple post-estimation contrasts did not show any significant differences in FIT-60 scores between ACT-DL and TAU at any time point (see Table 3).

**Table 3.**
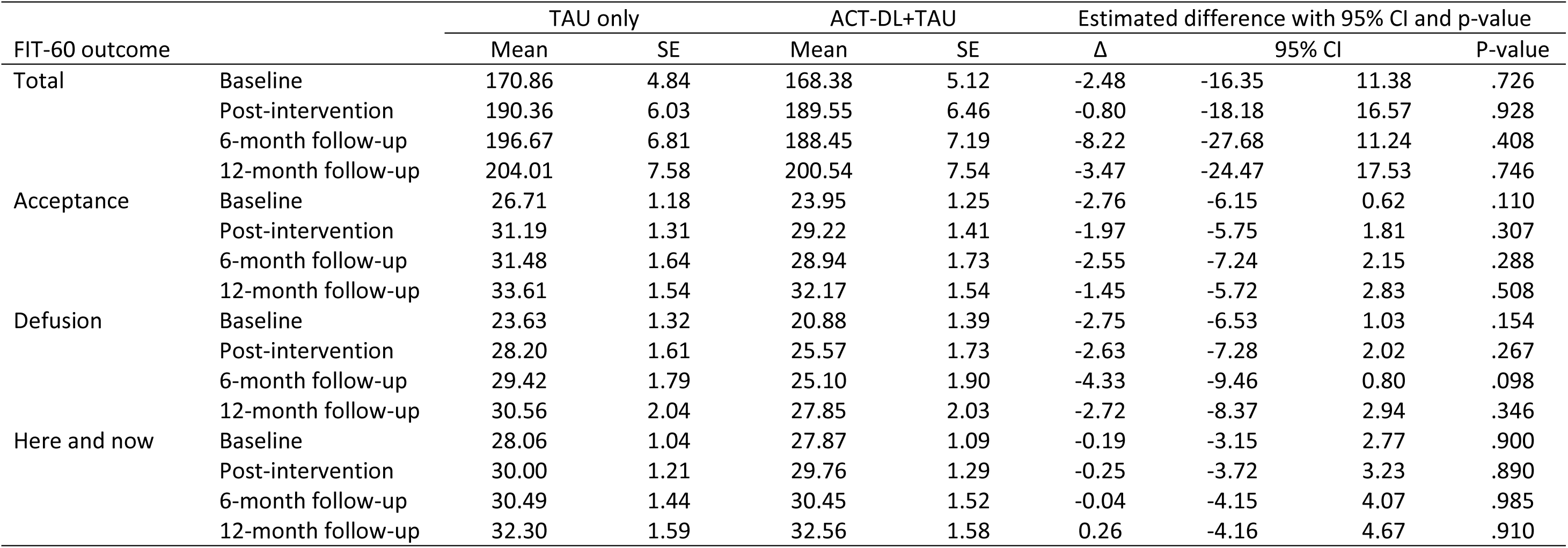
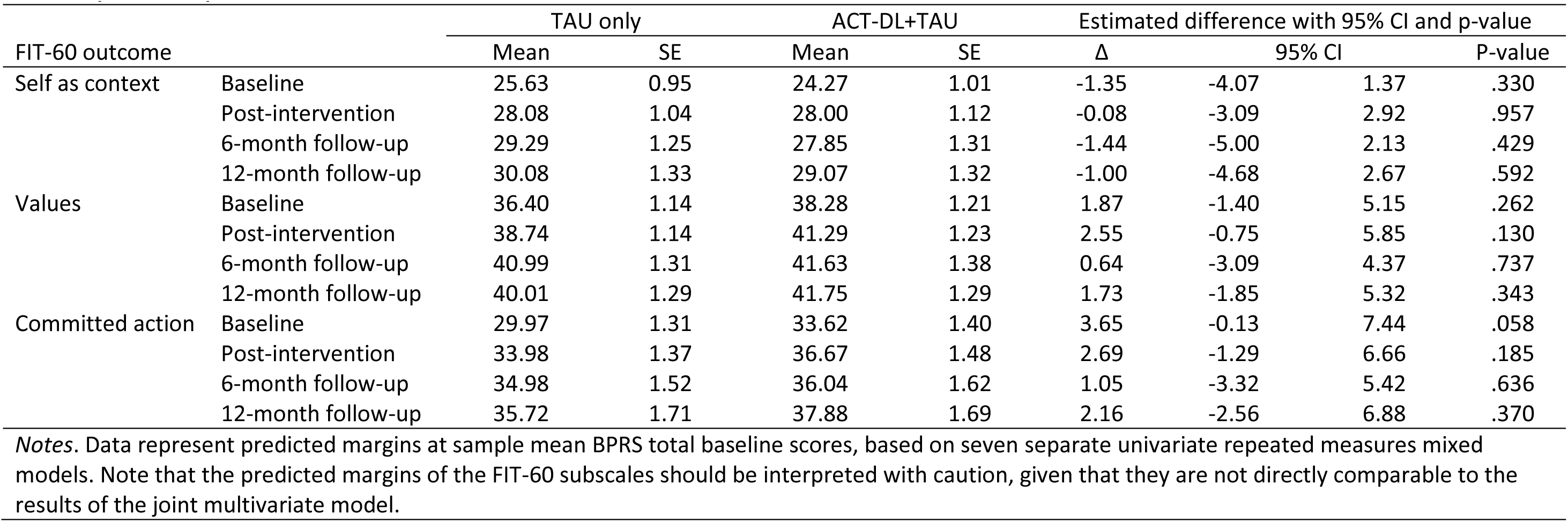
Predicted margins, SEs, and estimated differences between treatment conditions for global psychological flexibility and its six sub-processes.

#### 3.1.2 Daily life psychological flexibility

As is shown in Table 4 and Figure 4, the results of the three repeated measures mixed-effects models provided partial support for the hypothesis that reveal that individuals who followed ACT-DL+TAU showed increased improvement of daily life openness as measured with ESM outcomes, in comparison to those in the TAU condition.

**Figure 4.**
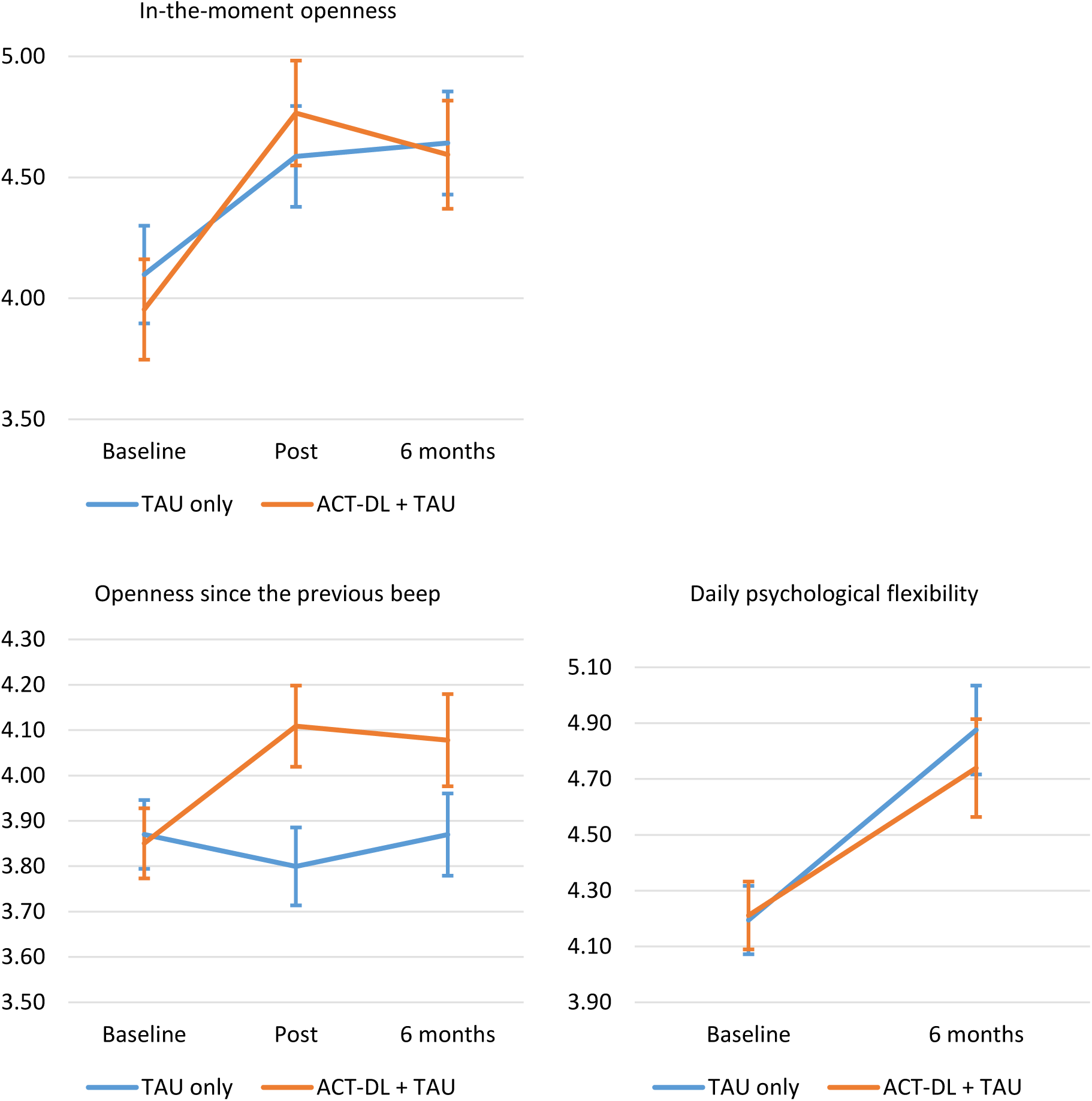
Daily life psychological flexibility at baseline, post-intervention, and 6-month follow-up. Data for daily psychological flexibility at post-intervention were unavailable due to technical issues.

**Table 4.**
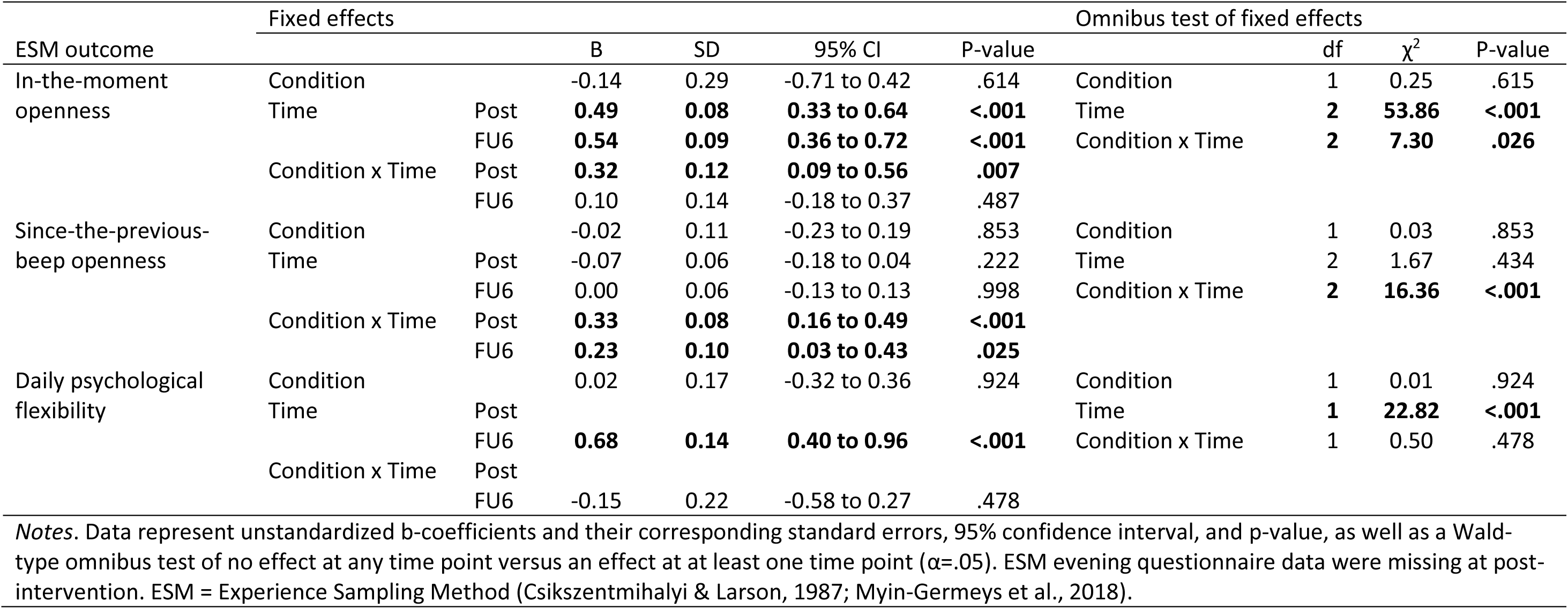
Results of repeated measures mixed-effects models of treatment effects on daily life psychological flexibility processes.

A significant Condition x Time interaction effect (P-value=.026) was found for in-the-moment openness, with time-specific contrasts indicating that individuals in ACT-DL+TAU, on average, were less inclined to avoid their negative feelings at post-intervention (P-value=.007), reflected by a steeper increase from baseline to post-intervention in in-the-moment openness as visualised in Figure 4. However, as shown in Table 5, these scores were not significantly different from each other at any other time point in the study as tested with simple post-hoc contrasts. While the increase in these scores was thus largest for ACT-DL individuals at post-intervention, a main effect of Time (P-value<.001) did indicate an increase in in-the-moment openness for the full sample, at both post-intervention (P-value <.001) and 6-month follow-up (P-value<.001).

**Table 5.**
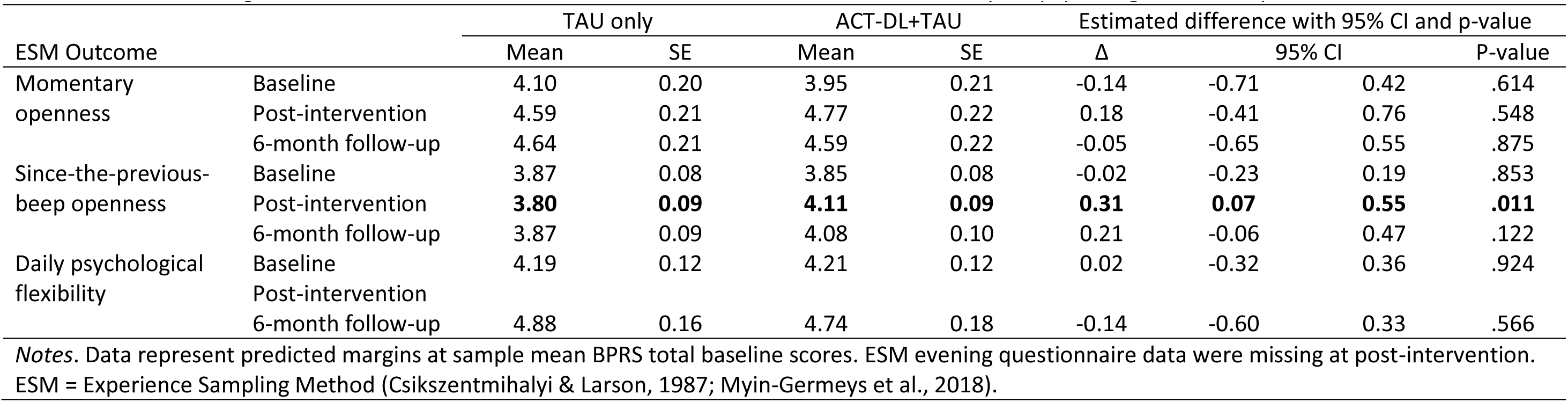
Predicted margins, SEs, and estimated differences between treatment conditions for daily life psychological flexibility.

Since-the-previous-beep openness scores also increased more in the ACT-D+TAU condition as opposed to TAU (P-value<.001) and did so at both post-intervention (P-value<.001) and 6-month follow-up (P-value=.025). Post-hoc simple contrasts equally showed significant differences in scores at post-intervention favoring ACT-DL+TAU (P-value=.011). The main effect of Time disappeared when introducing the Condition x Time interaction term, which suggests that the significant increases in these scores in the ACT-DL+TAU condition explained this effect.

Our hypothesis of a steeper increase in ACT-DL+TAU versus TAU was unconfirmed for daily PF scores. Yet, a significant main effect of Time was found at 6-month follow-up (P-value<.001). Finally, post-hoc contrasts showed that there were no significant differences between conditions at any time point for this outcome.

[Insert Tables 4-5 / Figure 4 here]

### 3.2 The moderating role of the therapeutic working alliance

#### 3.2.1 WAI-SF as a moderator of global psychological flexibility

As can be seen in Table 6 and Figure 5, the results of our moderation analysis did not provide support for the hypothesis that better client-perceived, or therapist-perceived WAI-SF would lead to larger increases in mean levels of FIT-60 total scores.

**Figure 5.**
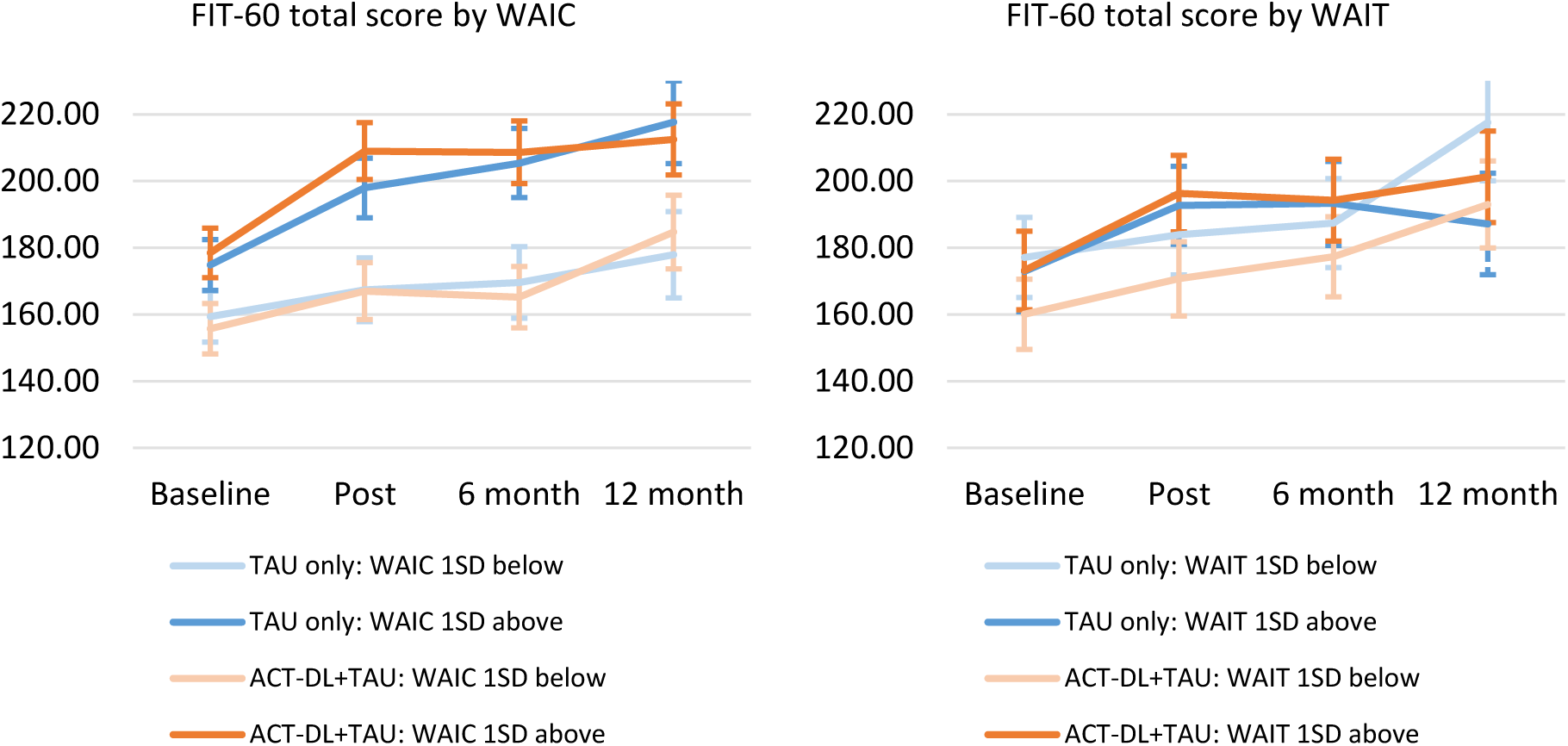
Global psychological flexibility by client-perceived working alliance (left) and therapist-perceived working alliance (right).

**Table 6.**
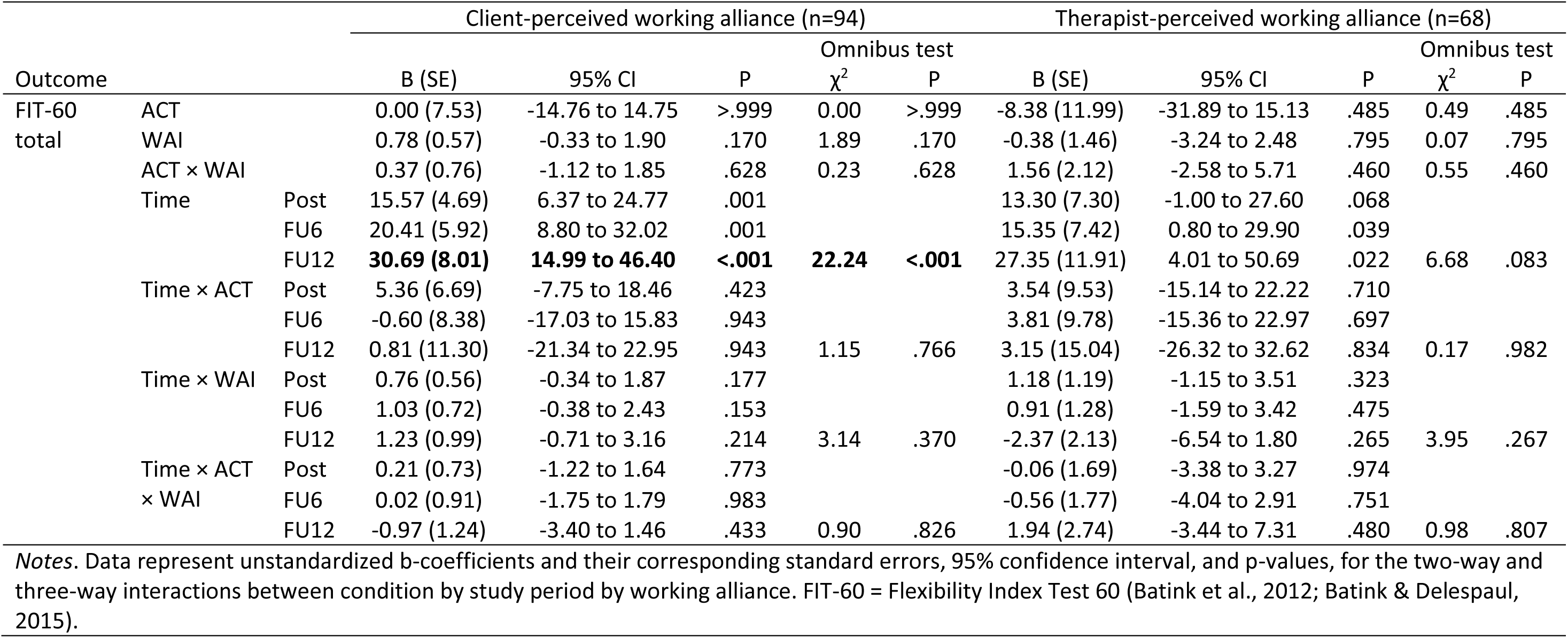
Treatment effect on global psychological flexibility by (a) client-perceived and (b) therapist-perceived working alliance.

#### 3.2.2 WAI-SF as a moderator of daily life psychological flexibility

Table 7 and Figure 6 show the moderating effects of client-perceived (left) and therapist-perceived (right) WAI-SF on ESM outcomes.

**Figure 6.**
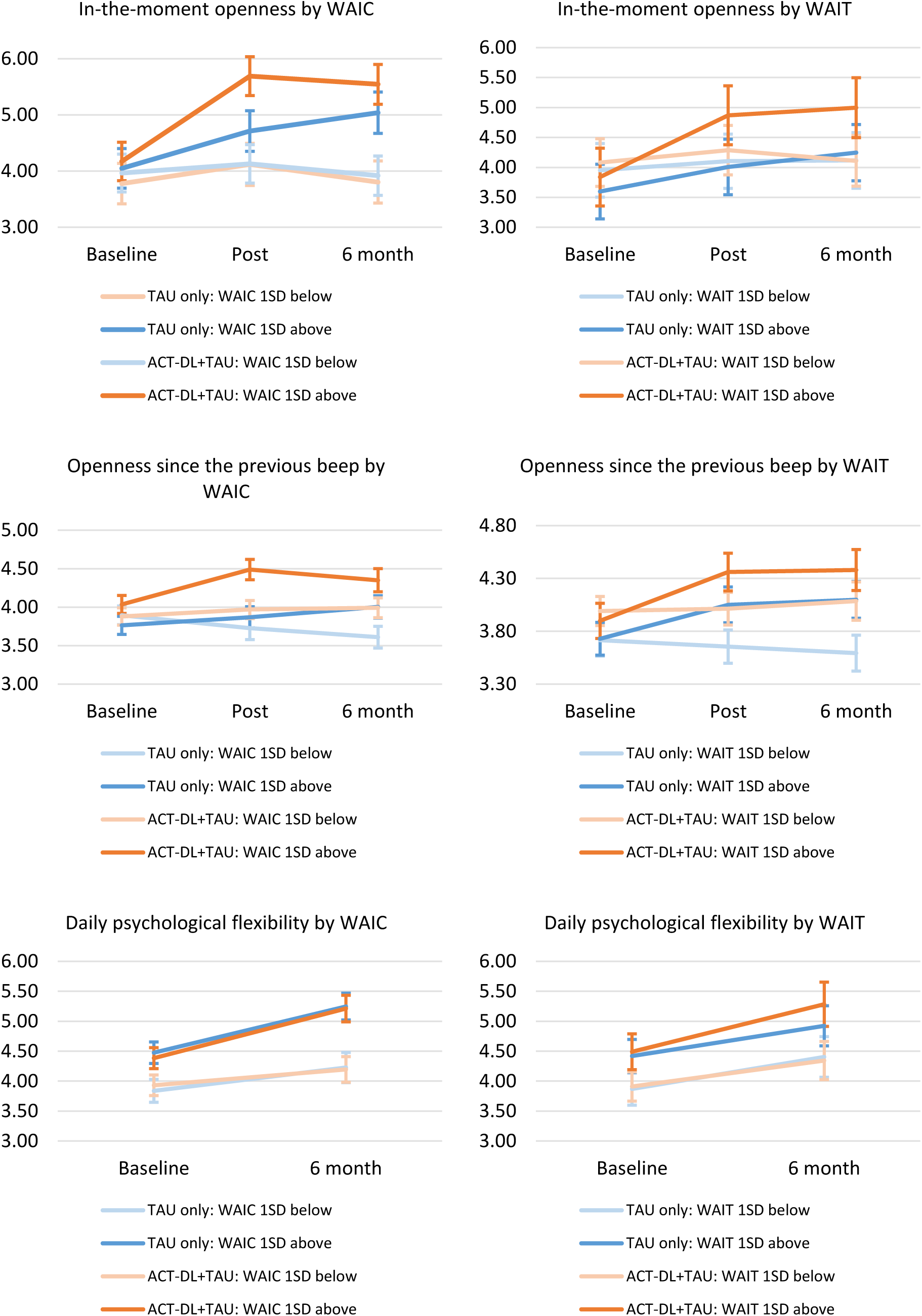
Daily life psychological flexibility by client-perceived working alliance (left) and therapist-perceived working alliance (right).

**Table 7.**
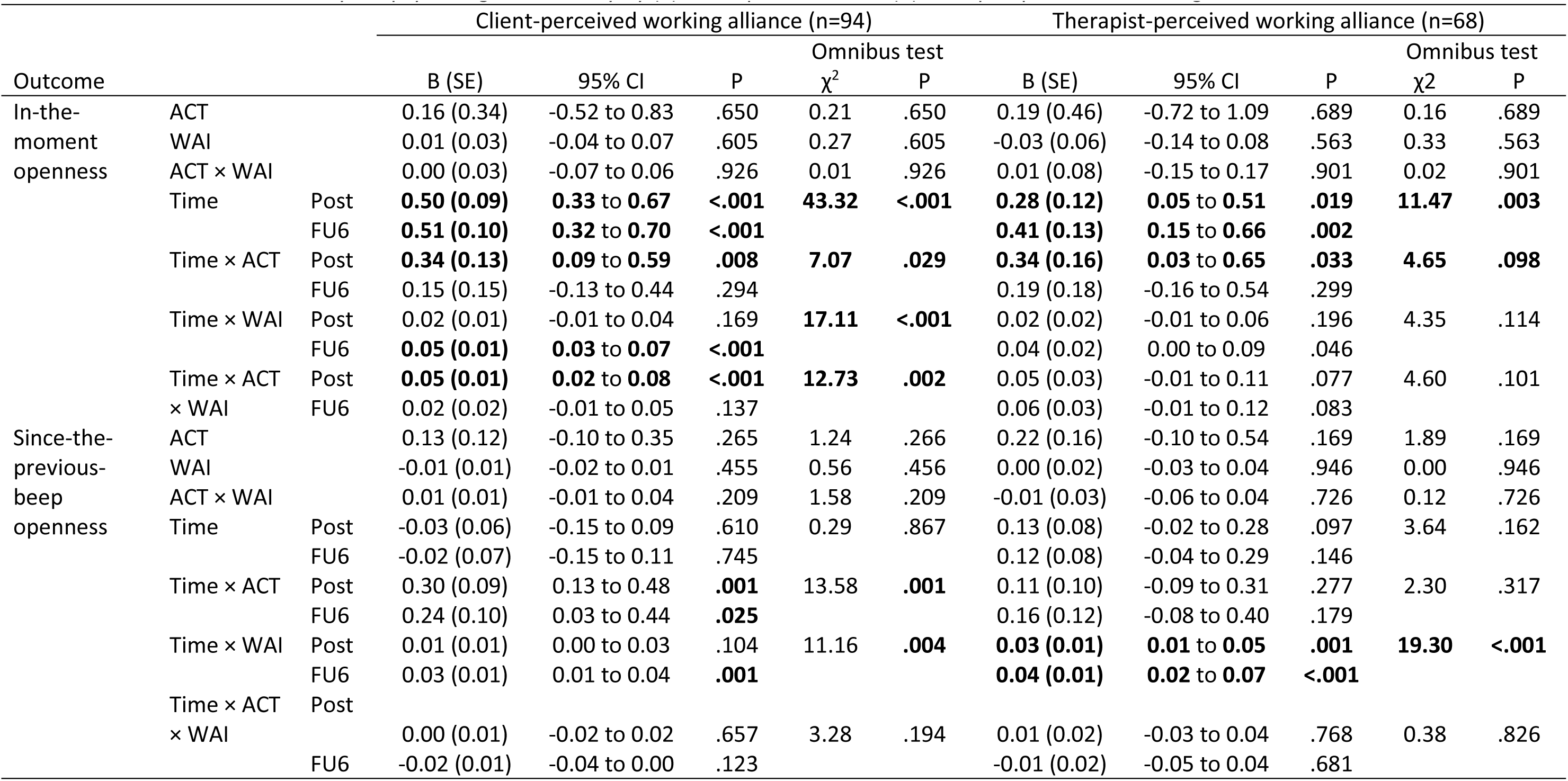
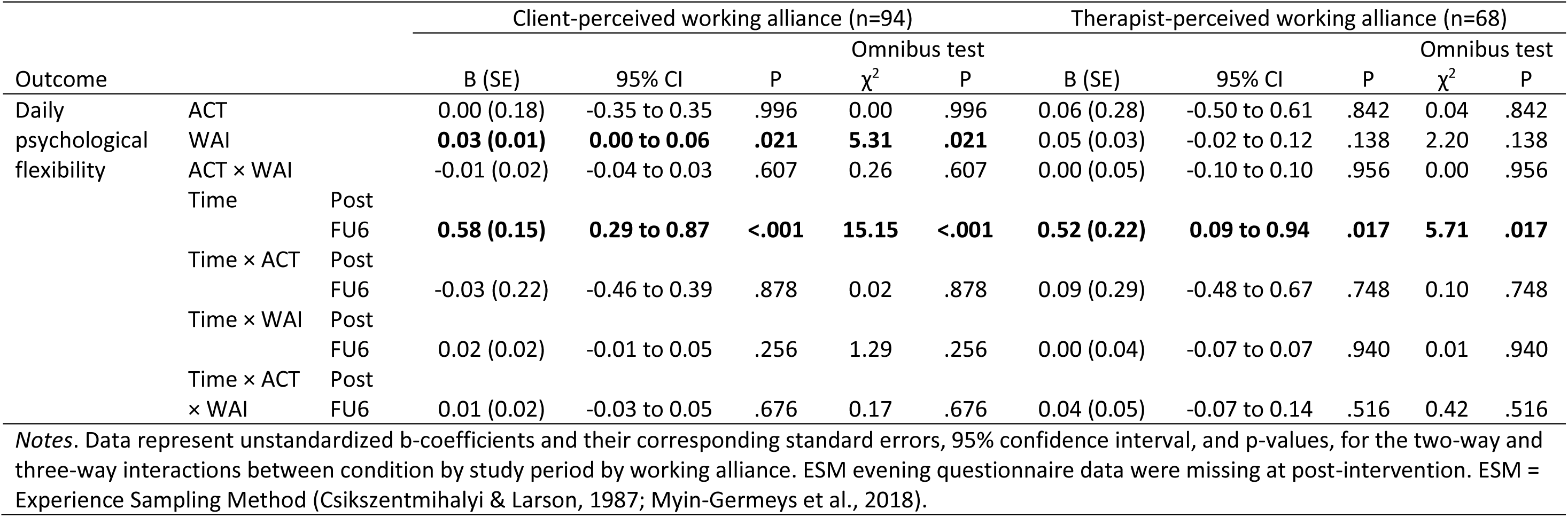
Treatment effect on daily life psychological flexibility by (a) client-perceived and (b) therapist-perceived working alliance.

Client-perceived working alliance moderated treatment effects of ACT-DL+TAU versus TAU on in-the-moment openness (P-value=.002), confirming our hypothesis that the largest increases in in-the-moment openness would be found for ACT-DL individuals who reported a better therapeutic working alliance and smallest increases for TAU individuals who reported worse working alliance, at least at post-intervention (P-value<.001). At the same time, there was a significant Time x WAI effect at 6-month follow-up, (P-value<.001), indicating that the increase in this ESM outcome from baseline to 6-month follow-up was steeper for those individuals who reported a better therapeutic working alliance in both conditions (see Table 7 and Figure 5). Therapist-perceived WAI-SF scores on the other hand did not moderate treatment effects on in-the-moment openness.

Furthermore, both client-perceived (P-value=.004) and therapist-perceived (P-value<.001) working alliance moderated the change in since-the-previous-beep openness in both conditions, from baseline to 6-month follow-up (P-value=.001) for the former, and from baseline to both post-intervention (P-value=.001) and 6-month follow-up (P-value<.001) for the latter.

Finally, a main effect of client-perceived WAI-SF was found on daily psychological flexibility scores (P-value=.021), indicating that those individuals who reported a better working alliance, also reported better scores on this outcome in general.

### 3.3 Service use checklist analysis (post-hoc analysis)

Analysis of our service use checklist (n=65) shows that 89% of the participants who were randomized to TAU did get some form of therapy at post-intervention, 81% at 6-month follow-up, and 89% at 12-month follow-up. Further inspection of this checklist informed us that TAU most commonly consisted of (various forms of) cognitive behavior therapy (CBT). Twenty-four participants received structured CBT for UHR or FEP (CBTp) (van der Gaag et al., 2012). Mindfulness-based group sessions, EMDR, and individual counseling sessions were also reported. Please refer to Table 8 for more details.

**Table 8.**
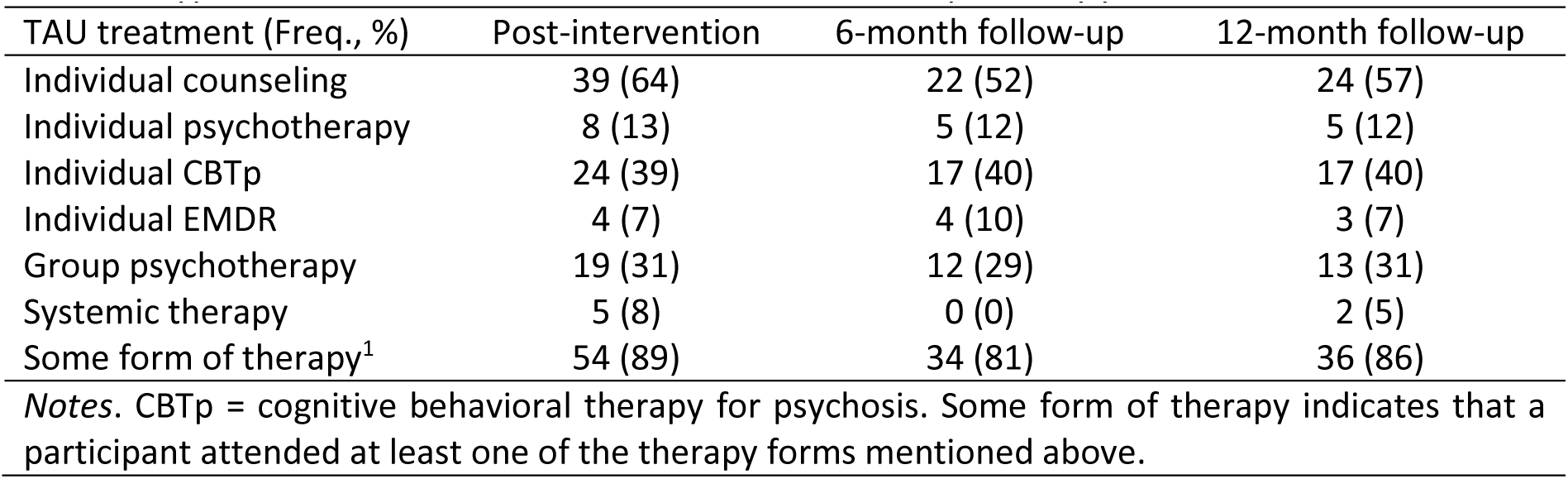
Types of interventions within the control condition per study period.

## 4 Discussion

This study was the first to investigate the effect of ACT-DL+TAU in comparison to TAU on both a global as well as daily life measures of psychological flexibility, including a long-term follow-up in a large sample size. We found that global and daily PF and its related sub-processes measured with the FIT-60 did improve to an equal extent in both the ACT-DL+TAU and TAU condition. In contrast, in-the-moment openness and since-the-previous-beep openness did increase more in ACT-DL+TAU in comparison to TAU (at post-intervention for in-the-moment openness, and at post-intervention and 6-month follow-up for since-the-previous-beep openness). Those participants in the ACT-DL+TAU condition reporting the best client-perceived working alliance showed the largest increases in in-the-moment openness, at least at post-intervention. In both treatment conditions, those participants with the best therapist-perceived working alliance had the largest increases in since-the-previous-beep openness at both post-intervention and 6-month follow-up.

### 4.1 Global psychological flexibility

Although in line with results from some prior clinical trials (Shawyer et al., 2012, 2017; White et al., 2011), the lack of a between-condition difference on PF improvement contrasts with other trials that found significant effects favoring ACT over TAU on acceptance (Gumley et al., 2017; Spidel et al., 2018) and mindfulness (Gumley et al., 2017; White et al., 2011). However, individuals in the ACT-DL+TAU condition improved significantly over time on all PF processes, and our hexaflex analysis suggest that these improvements were also clinically relevant when comparing scores to those based on a clinical population with heterogeneous psychopathology (Batink & Delespaul, 2015).

Whereas in a previous study from our group, ACT-DL failed to have significant effects on PF (van Aubel et al., 2020), in the current study, PF increased in both conditions, meaning that PF improved in TAU as well. We did not have full control over the specific therapeutic elements received in TAU and there might have been similar elements in both conditions, explaining the increase in PF scores in this condition. A majority of participants in the control condition received either individual or group counselling/psychotherapy, including various forms of CBT (e.g., schema therapy), as well as more recent forms of behavior therapy (e.g., mindfulness-based group intervention), which may explain the effect in the TAU condition. Indeed, ACT is similar to other (contextual) behavioral therapies in the sense that it incorporates experiential exercises to increase mindful awareness and acceptance, self-compassion, value clarification, and behavior activation (Hayes et al., 2011). Nonetheless, it is also argued to be specific in its effect on PF, given that all ACT-processes are integrated with a specific focus on the *willingness* to contact difficult experiences (acceptance) within the context of commitment to important personal values (Hayes et al., 2006). As such, the flexible use of the various coping skills to stay committed to personal values despite difficult experiences is ACT-specific and it remains unclear why there was no larger increase found in ACT-DL+TAU versus TAU on any of the FIT-60 subscales.

### 4.2 Momentary processes

Our findings on increased momentary openness in the ACT-DL+TAU condition are in line with results from another study that demonstrated significant improvements on momentary acceptance, defusion, valued living, and mindfulness (Levin et al., 2017). It is of note that, although of similar magnitude to previous ESM studies (Klippel et al., 2017), the observed increases in ESM openness measures were relatively small. However, in ESM studies, even small increases in therapeutic effects may be relevant given that they occur in the flow of daily life, and, over time, may accumulate into larger relevant effects (Kramer et al., 2014). In addition, the significant interaction effect did not hold for daily psychological flexibility as measured with ESM evening questionnaires. A limitation of the current study was that we did not have daily ESM data available at post-intervention due to technical issues. One possible explanation for this null-result is therefore that, similar to in-the-moment openness, the effect occurred at post-intervention but did not persist until the 6-month follow-up.

Yet, the effects for daily life openness are promising, given that multiple studies showed that low openness towards aversive mood states and cognitions is associated with reduced wellbeing and other negative outcomes including social anxiety and paranoia (Hershenberg et al., 2017; Levin et al., 2018; Machell et al., 2015; O’Toole et al., 2017; Udachina et al., 2009, 2014; Wenze et al., 2018). That is, low openness towards unwanted experiences may paradoxically exhaust the cognitive and emotional strategies needed to engage effectively in valued activities or important social interactions (Kashdan et al., 2014). This may be underscored by the fact that we found significant treatment effects in the INTERACT study on functioning, negative symptoms, and momentary symptom-related distress (Myin-Germeys et al., 2021). A next step will now be to test whether momentary openness is the actual process of change within ACT-DL making use of multilevel mediation models.

The results on daily life openness are in contrast with the results on global measures. A similar contrast emerged in our efficacy data (Myin-Germeys et al., 2021), where we found a significant effect on momentary psychotic distress, but not general psychotic distress as measured with a clinical interview. These results thus seem to suggest that the assessment of psychologically flexible behavior at the micro-level may be much more sensitive to subtle therapy-induced changes than assessment of more abstract metacognitive conceptualizations of this behavior at the macro-level. Nonetheless, it remains poorly understood how to operationalize psychologically flexible behavior at the micro-level with most studies to date, including ours, focusing exclusively on items assessing momentary (Udachina et al., 2009, 2014) and daily (Kashdan et al., 2014) acceptance, while neglecting other important ACT processes as well as their integration into PF. Future work is now necessary to capture this concept as a state in individuals’ daily lives.

### 4.3 Therapeutic alliance

Although inconsistent across the two measures of momentary openness and across time points, our results showing that a better client-perceived therapeutic alliance amplified effects on momentary openness, especially in ACT-DL+TAU at 6-month follow-up, add to previous studies that have repeatedly linked the quality of the therapeutic relationship to higher therapy engagement as well as improvements in overall psychotic, global, and negative symptoms (Bourke et al., 2021; Browne et al., 2021; Shattock et al., 2018). Our results underscore that a positively perceived working alliance is important for changing both therapeutic processes as well as outcomes, at least in daily life. This may even be more so due to the nature of the therapeutic relationship within ACT, where the various ACT processes need to become evident in the relationship between therapist and client itself. That is, ACT therapists themselves need to be vulnerable and open towards difficult experiences within the therapeutic process (Vilardaga & Hayes, 2009; K. G. Wilson & Sandoz, 2010).

### 4.4 Limitations

The current study needs to be interpreted within the context of some limitations. A first limitation was that we did not have data available on client-perceived working alliance for all participants. Although speculative, this could mean that we excluded those participants from our analyses that were the least engaged within their therapeutic trajectory, potentially biasing our moderation analysis. Second, the alliance was measured at post-intervention. As such, we cannot preclude that the therapeutic alliance was not a moderator of improvements in PF, but that is was actually an outcome of improved PF in itself, given that previous process literature has shown that improvements in symptoms throughout therapy may lead to a better perceived therapeutic alliance (Ardito & Rabellino, 2011; Folmo et al., 2021). Third, the operationalization of our daily life PF measures consisted only of a limited number of ESM items, and our momentary measures focused solely on the open ACT processes. Including the aware and active components of the PF model on the momentary level would have provided us with a more complete picture on daily life PF. Fourth, we cannot rule out that improved global PF within the ACT-DL condition was a result of increased familiarity with the concepts within the FIT-60 questionnaires thereby increasing social desirability to rate oneself higher on the questionnaire items, rather than the result of improvements in actual PF. Nevertheless, our finding that global PF improved to an equal extent in both conditions seems to point into the direction of an actual increase in PF over time.

### 4.5 Conclusions

In conclusion, while we did not find any significant treatment differences on global psychological flexibility, we did find improvements in PF in both conditions, as well as treatment effects favoring ACT-DL on two momentary measures. In addition, our results provide further support for the importance of a good therapeutic relationship between client and therapist. The therapeutic effects were visible there where we provided therapeutic support by making use of our ACT-DL EMI, that is, in the daily lives of our participants. Next steps are now to unravel the mediating role of daily life openness of ACT-DL on important therapeutic targets in early psychosis. In addition, future work is now needed to improve our understanding of daily life PF and how to operationalize it.

## Data Availability

Deidentified data are available upon request through a data access system, Data cuRation for OPen Science (DROPS), administered via REDCap at the Center for Contextual Psychiatry, KU Leuven. Interested researchers can submit an abstract, which is subject to review by the research team to ensure there is no overlap with existing projects. Following abstract approval, a variable access request is submitted and researchers are required to postregister their analysis plan. A dataset containing only variables required for the proposed analysis is then released to the researchers by a data manager, along with a time- and date-stamped receipt of data access.

https://osf.io/jzyp2/?view_only=9672553dd8fd461ebe10e592ed42c707

## Acknowledgements

We thank all the participating health services in Amsterdam (Academic Medical Centre, Arkin Basis GGZ), The Hague (Parnassia, PsyQ), Maastricht/Eindhoven (Mondriaan, Virenze, GGZE), Flemish-Brabant (UPC KU Leuven, VDIP Antwerp, Sint-Annendael, PCM Mortsel), and East/West Flanders (OLV Brugge, Karus Melle, VDIP Sint Niklaas). We thank all research coordinators (Silke Apers, Nele Volbragt, Wendy Beuken), research assistants (Dieuwke Siegmann, Davinia Verhoeven, Anna de Zwart, Iris de Wit, Lore Depraetere, Tessa Biesemans, Lotte Hendriks), and data managers (Martien Wampers, Jolien Bynens) past and present who were involved in the INTERACT trial. We also like to thank all individuals who participated in the study and were essential for its successful completion.

## Footnotes

a Given the multifinality of psychiatric outcomes in UHR individuals (Beck et al., 2019; Lin et al., 2015), along with the finding that transition rates to a psychotic disorder were lower in more recent samples (Hartmann et al., 2016; Nelson et al., 2016), the last couple of years the term clinical high risk (CHR) has also been used to refer to individuals in the UHR group (McGorry & Mei, 2018; Nelson & McGorry, 2020). We chose the term UHR for consistency with the INTERACT study protocol (Reininghaus et al., 2019) and main outcome paper (Myin-Germeys et al., 2021).

b The current study was post-registered on the Open Science Framework (https://osf.io/), meaning that we registered our analysis plan after we had collected data, however before we had any access to the data. The main post-registration for the INTERACT study is available here https://osf.io/du2bn/?view_only=ec22ed02651441349e1bb1242cfc712c. The post-registration for the current study is embedded as a file within the main registration and can be accessed here https://osf.io/jzyp2/?view_only=9672553dd8fd461ebe10e592ed42c707.

