## Supplement for "Psychological flexibility and the moderating role of the therapeutic working alliance in Acceptance and Commitment Therapy in Daily Life (ACT-DL) in an early psychosis sample"

### Supplementary materials

*Supplement 1. ESM compliance and validity of the daily life measures*

Compliance to the ESM beep questionnaires started at 39% at baseline, and dropped to approximately 30% at post-intervention and 6-month follow-up. Participants filled in on average 4 out of 6 evening questionnaires at baseline and at 6-month follow-up. ESM evening questionnaire data at post-intervention is missing for all participants due to technical errors.

To assess concurrent validity of global and daily life psychological flexibility, we calculated baseline Pearson-correlations between all person-mean ESM outcomes on the one hand and the baseline FIT-60 total and subscale scores on the other. In-the-moment openness correlated significantly (all $P<.001$) with the open (acceptance, defusion) and aware (self as context, mindfulness) FIT-60 subscales, with moderate to large correlation coefficients between .39 (self as context) and .58 (defusion). Since-the-last-beep openness correlated moderately with total psychological flexibility ($r.41$; $p=.011$), acceptance ($r = .30$; $P=.014$), defusion ($r = .37$; $p<.001$), and mindfulness ($r= .31$; $P<.001$). Daily psychological flexibility correlated significantly with all ACT processes (all $P<.01$, $r=.36$ to .60). These correlations thus provided support for the convergent validity of our daily life psychological flexibility measures with retrospective psychological flexibility (see Supplementary Table 1).

### Supplementary tables

**Supplementary Table 1.**

Correlations between daily life and global psychological flexibility measures.

| | In-the-moment openness | | Since-the-last-beep openness | | Daily psychological flexibility | |
|---|---|---|---|---|---|---|
| Measure | Corr. | P-value | Corr. | P-value | Corr. | P-value |
| Global PF | 0.54 | <.001 | 0.31 | .011 | 0.59 | <.001 |
| Acceptance | 0.53 | <.001 | 0.30 | .014 | 0.36 | .001 |
| Defusion | 0.58 | <.001 | 0.37 | <.001 | 0.39 | <.001 |
| Mindfulness | 0.54 | <.001 | 0.31 | .009 | 0.44 | <.001 |
| Self as context | 0.39 | <.001 | 0.15 | >.999 | 0.43 | <.001 |
| Values | 0.21 | .360 | 0.03 | >.999 | 0.60 | <.001 |
| Committed action | 0.16 | >.999 | 0.13 | >.999 | 0.52 | <.001 |

Notes. PF = Psychological flexibility. Data represent Bonferroni corrected pairwise correlations and p-values. Corr. = correlation.

### Supplementary figures

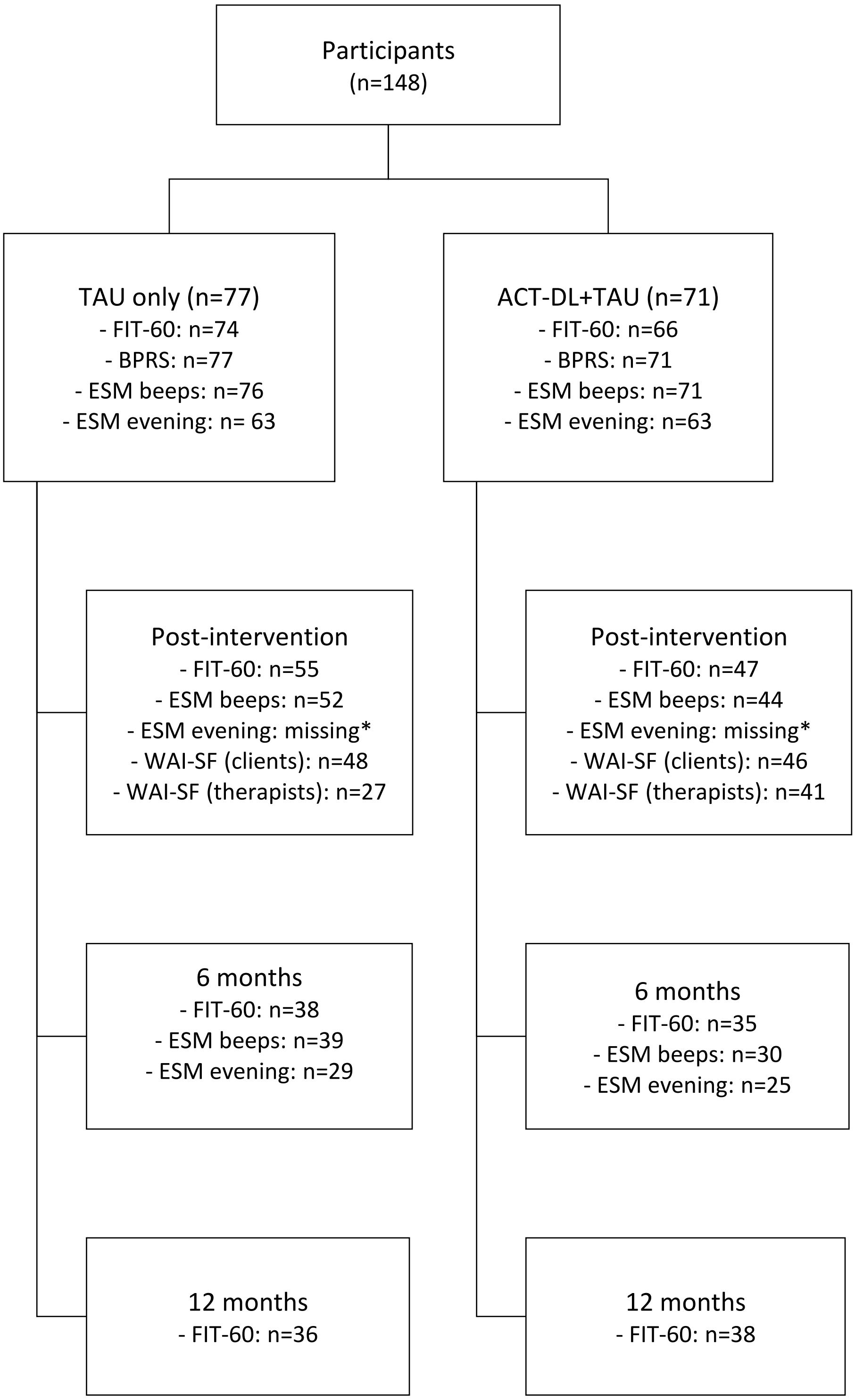


*Supplementary Figure 1*. Study flowchart for ACT-DL processes. * ESM evening questionnaire data at post-intervention was missing due to technical issues.
